# Multimorbidity patterns among COVID-19 deaths: considerations for a better medical practice

**DOI:** 10.1101/2020.07.28.20163816

**Authors:** Julián A. Fernández-Niño, John A. Guerra-Gómez, Alvaro J. Idrovo

**Author notes:** **Correspondence:** Julián A. Fernández-Niño. Public Health Department, Universidad del Norte, Barranquilla, Colombia.

## Abstract

Medical care of individuals diagnosed with severe COVID-19 is complex, especially when patients are older adults with multimorbidity. The objective of this study was to describe patterns of multimorbidity among fatal cases of COVID-19. Data of Colombian confirmed deaths of COVID-19 until June 11, 2020, were included in this analysis (1488 deaths). Relationships between COVID-19, combinations of health conditions and age were explored using locally weighted polynomial regressions. Some multimorbidity patterns increase probability of death among older individuals, whereas other patterns are not age-related, or decreases the probability of death among older people. Consider multimorbidity in the medical management of COVID-19 patients is important to determine the more adequate medical interventions. In addition to the co-occurrence of COVID-19 with diseases of high prevalence in the world, in Colombia there are cases more complex with COVID-19 co-occur with endemic and orphan tropical diseases. In these cases, although its occurrence may be low, clinical management requires adjusting to its complex clinical condition.

## Introduction

Currently, the practice of medicine is complex, and it is even more so when faced with a new infection such as COVID-19.^1^ Scientific knowledge provides guidelines for having an effective medicine, but when people have chronic diseases, more than one disease (co-occurrence), and receiving several medicines, clinical practice becomes an even bigger challenge, especially when these chronic diseases do not receive adequate care.^2^ In addition, conditions such as physical disability, mostly also as a consequence of these chronic diseases, could also increase vulnerability to complications from other different diseases.

Evidence show that multimorbidity is one of the clinical characteristics that most complicate the care of infected people,^3^ which is clearly true for COVID-19 as well. Valderas et al.^4^ defined multimorbidity as the “presence of multiple diseases in one individual”, where comorbidity is just one of the possible forms of multimorbidity. For these authors comorbidity is the “presence of additional diseases in relation to an index disease in one individual”, therefore, defining the co-occurrence of various pre-existing diseases with COVID-19 infection is not necessarily comorbidity.

Multimorbidity among individuals with COVID-19 was described early among the first reported patients in the specialized literature,^5^ and it is a frequent finding in published studies.^6,7^ However, the term comorbidity continues to be used incorrectly in most published studies. A critical reading of previous studies allows us to identify that, for the most of these. the objective was to explore the effect of single diseases on the clinical severity or the risk of fatality among patients with diagnosis of COVID-19. This leaves out more complex approaches such as those that configure vulnerability of older adults as a construct beyond the effect of single diseases or their simple sum. Therefore, it is necessary to study multimorbidity as a multidimensional condition, which incorporates the joint presence and the interactions of all the health conditions. A deeper analysis should emphasize the type of potential relationships between health conditions, whether they are under medical control or not, as well as other sources of vulnerability such as disability, functional dependence on other people, the presence of geriatric syndromes or even the social and economic vulnerabilities, among others. Understanding the multimorbidity associated with emerging diseases such as COVID-19 is important to classify patients and define differential medical management guidelines.^8^

In critical situations such as the COVID-19 pandemic, a good classification of infected people is important for improve their care. In addition, recognizing the multimorbidity as a multidimensional and complex condition will allow the development of more complex risk classifications, to identify subjects who need greater vigilance at the community and hospital level, or to formulate “telework” recommendations in certain occupations.

The wide variety of multimorbidity patterns includes the most frequent co-occurrences that were evident since the beginning of the pandemic, and the co-occurrences with less frequency that appear every time there are more cases, and perhaps it will include orphan diseases,^9,10^ and those of endemic foci.^11^ In these circumstances, low prevalence signifies more clinical complexity. In this study, we used Colombian data to describe patterns or multimorbidity among individuals diagnosed with COVID-19.

## Material and methods

Official nominal data of confirmed cases of COVID-19 for Colombia, with a cutoff date of June 11, 2020 throughout the country were included in this study. This data had a good performance according to an analysis based on Bedford’s law.^12^ By that date, 17790 cases and 1488 deaths by COVID-19 were reported. This database has information on each case, specifically: municipality and department of occurrence, date of onset of symptoms, date of diagnosis, date of report, sex, and age. For each dead individual, the presence or absence of a predetermined list of comorbidities is reported in the database: high blood pressure (HBP), diabetes mellitus, respiratory disease, cardiovascular disease, Chronic Kidney disease (CKD), cerebrovascular disease, smoking, cancer, thyroid disease, autoimmune disease, and HIV. This list includes nine of the top 10 chronic diseases identified in a systematic review of measurements of multimorbidity.^13^ Although smoking is not a chronic disease, it is a chronic health condition that has direct functional effects, beyond being a risk factor for other diseases, reason why it is incorporated as part of a broad definition of multimorbidity in this analysis.

Individuals reported as hospitalized or in the Intensive Care Unit (ICU) were not included, because they were in an intermediate stage of their medical treatment, and the definitive outcome was unknown (recovery or death). Information about chronic disease among mild and moderate cases of COVID-19 is not available by the time of this analysis. The database can be requested from the Colombian Ministry of Health and Health Protection.

### Statistical methods

First, the description of the prevalence of each disease among fatal cases was made individually, without considering the co-occurrence of other health conditions. These patterns were compared between age groups using *x*^*2*^ test. Subsequently, all possible disease dyads and triads were identified, characterizing multimorbidity. With this procedure were recognized the 10 most prevalent (>2%) multimorbidity patterns. For the estimation of prevalences, it was assumed that the individuals observed were a random sample of cases from the total population of COVID-19 cases that will be observed in Colombia at the end of the pandemic, which is reasonable given that the country has several weeks with sustained local transmission. Given this assumption, and that the prevalences are exceptionally low, exact 95% Clopper-Pearson confidence intervals were estimated.^14^ Cluster at the department level were considered to consider the spatial correlation of observations.

A locally weighted polynomial regression was fitted between each morbidity and age (continuous) to identify patterns of association. Finally, we proposed three simple qualitative models of multimorbidity for COVID-19 using configurations described by Valderas et al.^4^ The analyses were performed with the statistical software Stata 16 (Stata Corporation, USA). In addition, a real-time visualization was build based on the UpSet Visualization and the UpSet Altair implementation to display all the possible configurations of multimorbidity.

## Results

In this study, 1488 deaths were analyzed and 61% of them occurred in men. The mean of age of the fatal outcomes was 67.59 years (median: 69 years; percentiles: p_10_= 46, p_25_= 58, p_75_= 79, p_90_= 86 years) and only 27.19% of the deaths occurred among people under 60 years. In table 1 are the main health conditions accompanying COVID-19 infection, with their respective prevalences and 95% confidence intervals, according to the age groups (under 60, and 60 years and more). Prevalences of most of the health conditions are higher among individuals with 60 or more years. Exceptions were obesity, autoimmune disease, HIV, and diabetes + obesity, which were more frequent among individuals under 60 years. All these differences were significative (*p*<0.05). In figure 1 note that multimorbidity increases markedly its prevalence among those over 80 years.

**Table 1.** Prevalence of health conditions^¥^ among fatal cases of COVID-19 in Colombia.

| Only one condition co-occurrence | Under 60 years |  |  | 60 years and more |  |  |
| --- | --- | --- | --- | --- | --- | --- |
|  | Prevalence |  | 95% CI | Prevalence |  | 95% CI |
|  | (%) |  |  | (%) |  |  |
| HBP | 26.80 | 20.84 | 33.45 | 39.76 | 33.07 | 46.74 |
| Respiratory disease | 7.20 | 4.56 | 10.70 | 19.46 | 14.82 | 24.82 |
| Diabetes | 18.36 | 14.72 | 22.48 | 18.81 | 15.55 | 22.44 |
| Cardiovascular disease | 7.94 | 5.98 | 10.30 | 16.77 | 13.75 | 20.17 |
| Kidney disease | 5.71 | 4.58 | 7.57 | 11.86 | 10.20 | 13.69 |
| Obesity | 18.61 | 14.14 | 23.78 | 5.19 | 4.13 | 6.43 |
| Smoking | 3.97 | 1.87 | 7.31 | 5.19 | 3.61 | 7.20 |
| Stroke | 0.74 | 0.09 | 2.65 | 4.63 | 3.62 | 5.93 |
| Thyroid disease | 1.99 | 0.76 | 4.19 | 4.54 | 2.33 | 7.89 |
| Cancer | 2.48 | 1.07 | 4.87 | 3.89 | 2.38 | 5.97 |
| Autoimmune disease | 3.97 | 1.38 | 8.73 | 1.11 | 0.39 | 2.38 |
| HIV | 2.48 | 0.70 | 6.75 | 0.36 | 0.10 | 0.91 |
| **Two or more conditions co-occurrence** |  |  |  |  |  |  |
| HBP + Diabetes | 9.68 | 7.09 | 12.82 | 11.03 | 8.66 | 13.78 |
| HBP + Cardiovascular disease | 3.23 | 1.69 | 5.53 | 8.71 | 6.19 | 11.87 |
| HBP + Respiratory disease | 2.25 | 1.20 | 3.77 | 7.60 | 5.67 | 9.94 |
| HBP + Chronic Kidney Disease | 1.99 | 1.16 | 3.17 | 5.75 | 4.42 | 7.32 |
| Cardiovascular disease + Respiratory disease | 1.24 | 0.45 | 2.71 | 3.99 | 2.69 | 5.67 |
| Diabetes + Cardiovascular disease | 2.48 | 1.31 | 4.23 | 3.71 | 2.71 | 4.93 |
| HBP + Obesity | 5.21 | 3.18 | 7.99 | 2.87 | 2.16 | 3.74 |
| Diabetes + Cardiovascular disease + HBP | 1.49 | 0.54 | 3.25 | 2.50 | 1.76 | 3.45 |
| Smoking + Respiratory disease | 0.74 | 0.16 | 2.15 | 2.22 | 1.48 | 3.21 |
| Diabetes + Obesity | 4.96 | 3.46 | 6.86 | 1.67 | 0.88 | 2.87 |

\* Only dyads and triads with higher prevalences. HBP: High blood pressure.

**Figure 1.**
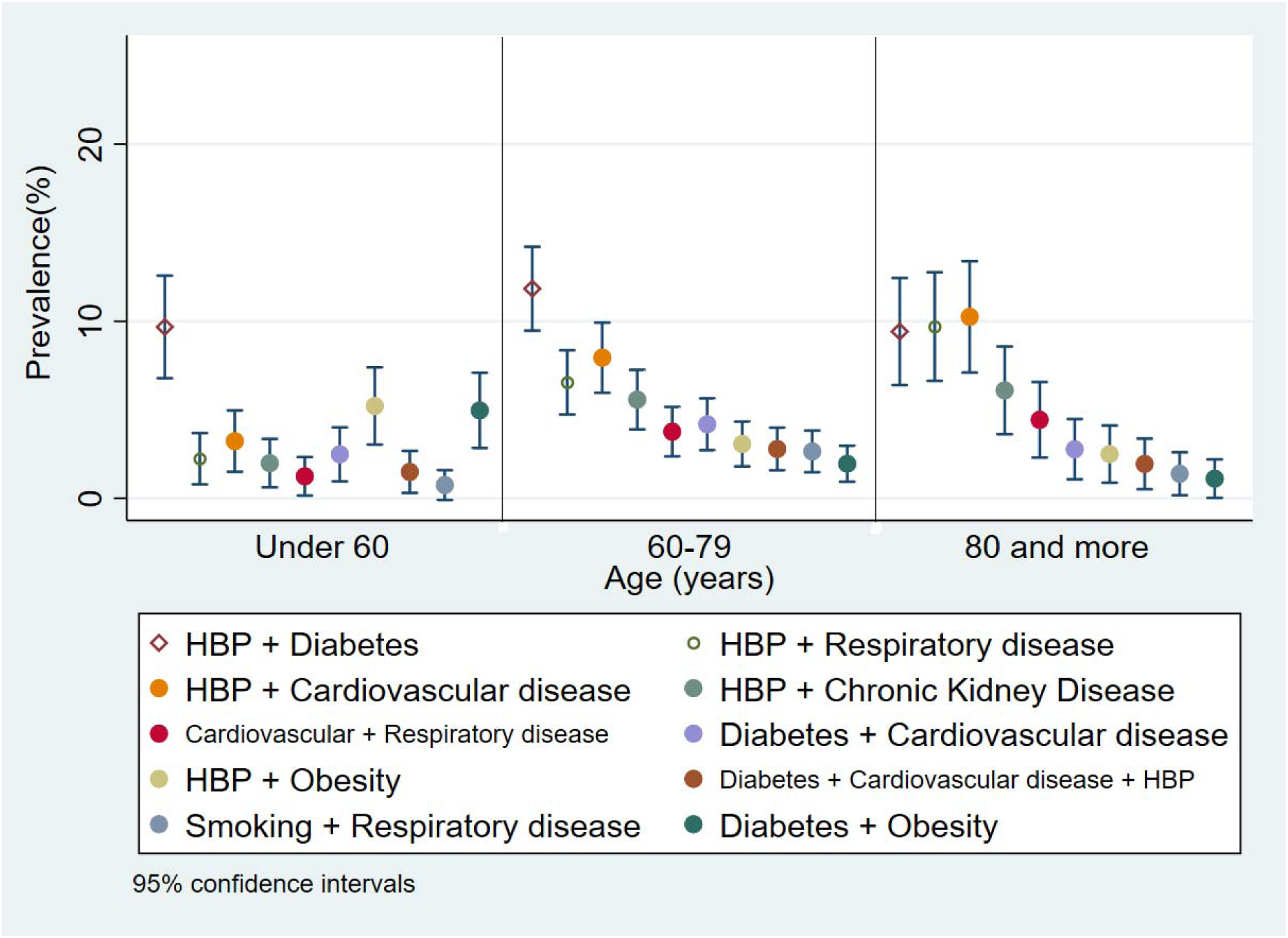
Prevalence of the 10 main dyads and triads identified among fatal cases of COVID-19 in Colombia, by age group.

In figure 2 are the probability of each morbidity according age, considered separately, estimated with locally weighted polynomial regressions. In general, there is a tendency to increase the probability of each morbidity with increasing age, and only high blood pressure, cardiovascular disease, and kidney chronic disease tend to be linear. In contrast, Obesity decreases the probability of occurrence with increasing age, whereas smoking, thyroid disease, cancer, autoimmune disease, and HIV have not a clear trend to increase or to decrease. In conclusion, among the fatal cases of COVID-19, the occurrence of multimorbidities is positively associated with age, as is known. One of the exceptions is obesity, which is more frequent in fatal young cases. Which is also shown by the fact that obesity has a higher occurrence in fatal cases in young people.

**Figure 2.**
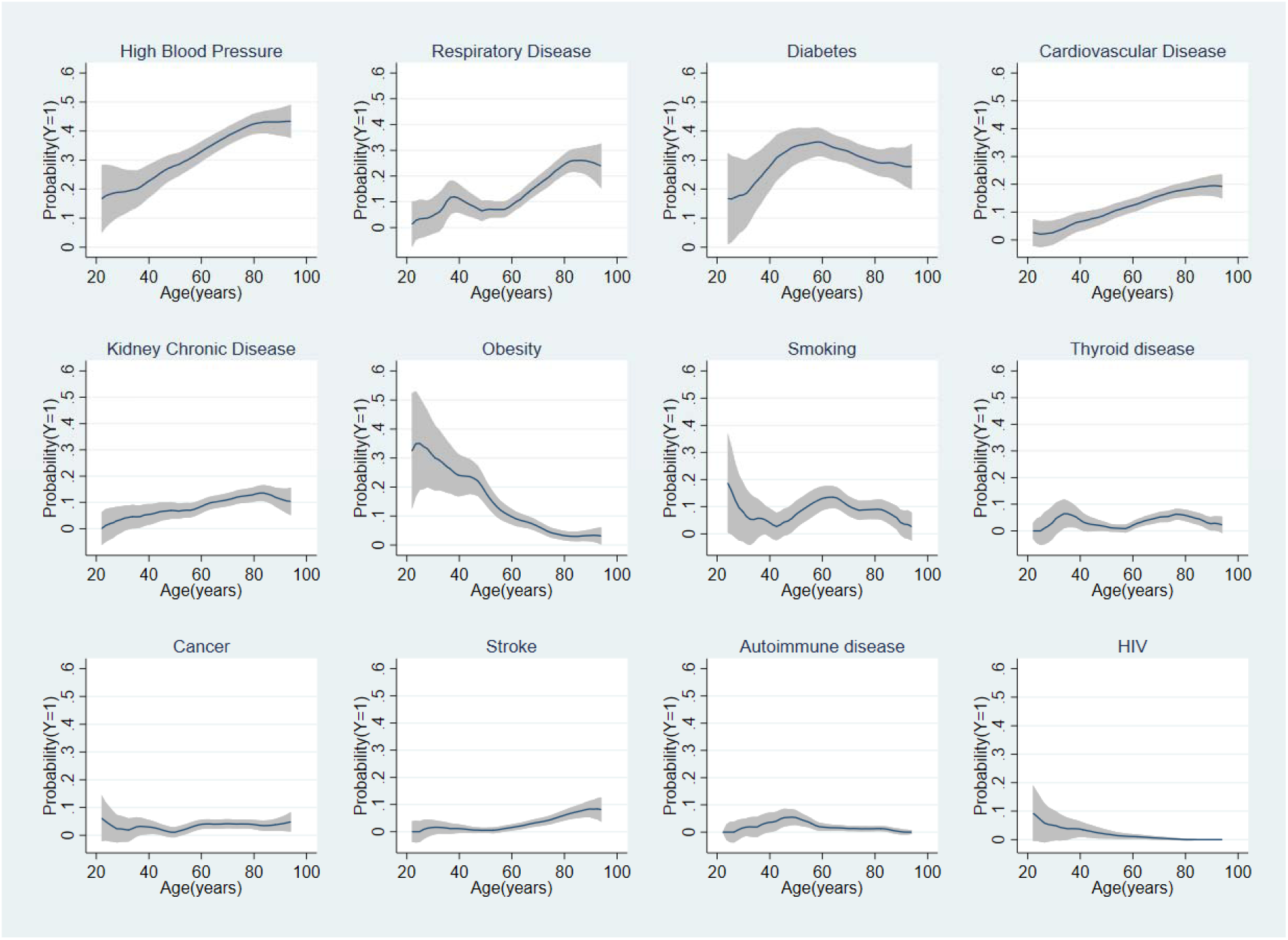
Locally weighted polynomial regressions between COVID-19, health condition and age, among fatal cases in Colombia. * * Only includes health conditions separately.

In figure 3 are the probability of each profile of multimorbidity according the 12 top complex health conditions (COVID-19 + two or more health conditions) according age, estimated with locally weighted polynomial regressions. In these cases, the relationships are heterogeneous and non-linearity is frequent. The COVID-19 + high blood pressure + diabetes pattern has an inverted U distribution and it is very important due to its relative high prevalence. It contrast with the linear relationship of COVID-19 + high blood pressure + respiratory disease. In summary, not all multimorbidity increases with age, as is commonly thought.

**Figure 3.**
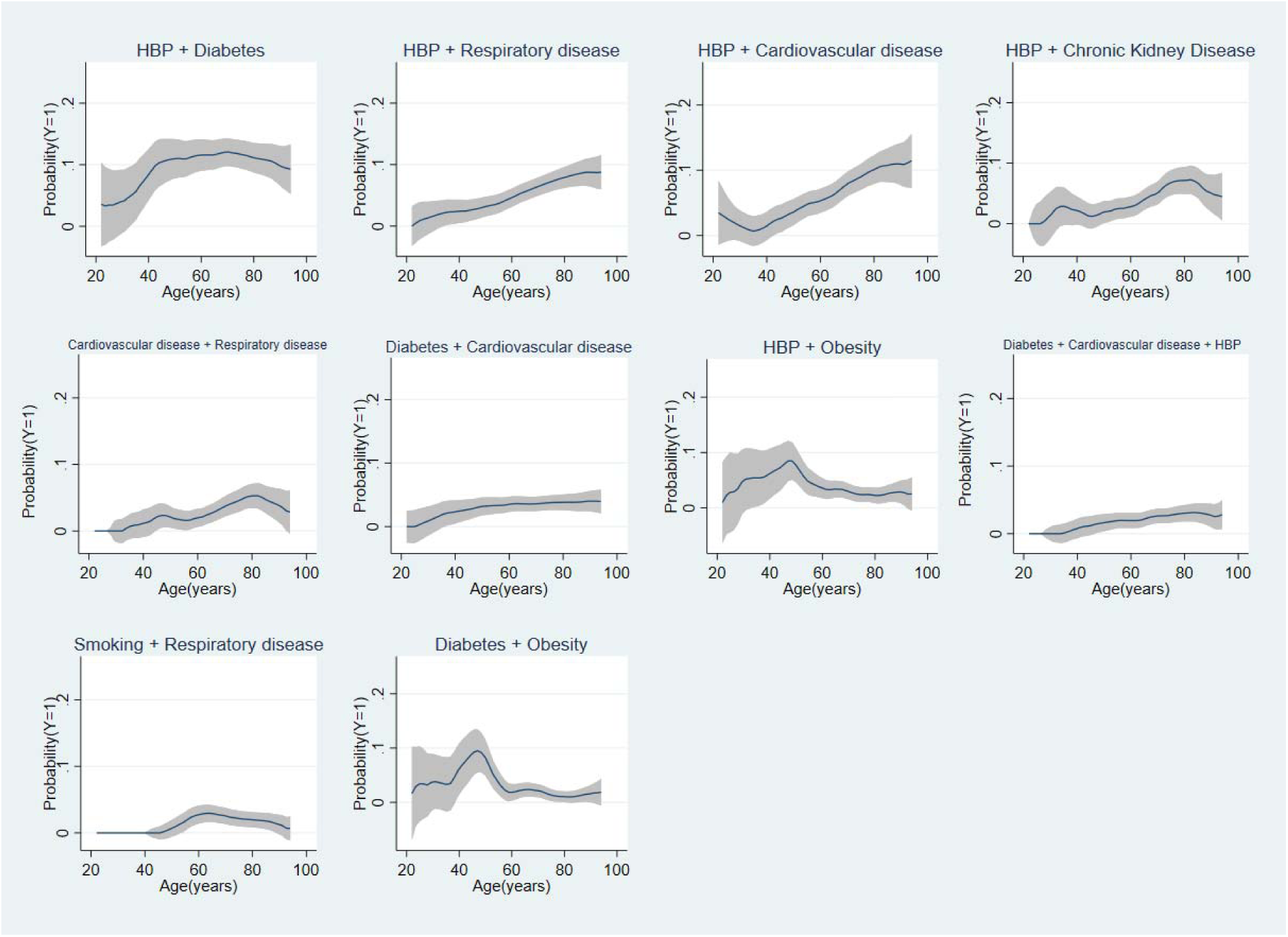
Locally weighted polynomial regressions between COVID-19, complex patterns of health conditions and age, among fatal cases in Colombia.* * Includes patterns of two or more health conditions.

## Discussion

Findings of this study indicate that multimorbidity is an important phenomenon to consider in the context of the COVID-19 pandemic. The co-occurrences of COVID-19 with different combinations of diseases including high blood pressure, diabetes mellitus, obesity, cardiovascular, respiratory, and CKD have a high frequency among individuals deceased by COVID-19 in Colombia and other countries.

The top six morbidities identified have a strong relationship with age (HBP, respiratory disease, diabetes, cardiovascular disease, CKD, and obesity). Most of these six associations were positive but not linear, except obesity and diabetes where the association decreases with age after a turning point. Given that these results are conditional on death, these relationships could mainly be explained by survivor bias, since diabetes and obesity are not only more prevalent in young adults, but also this in turn is the effect of higher mortality before 60 years of age. However, the importance of this finding is that it precisely identifies that diseases such as obesity and diabetes are more relevant in adults younger than 60 years as potential factors associated with COVID-19 fatality. Other diseases such as thyroid diseases, HIV, autoimmune diseases, stroke, and cancer seem to be important although not age-related. These diseases seem to be important regardless of age as factors associated with fatal outcomes of COVID-19 infection.

With the multimorbidity classification by Valderas et al^4^ is possible to identify that among individuals diagnosed with COVID-19 there are different types of multimorbidity. We propose that that there are least three possible models between COVID-19 and chronic diseases (figure 4). In the first model, chronic disease is an effect modifier of SARS-CoV-2 infection, increasing the risk of complications and severity. The second model incorporates the existence of common causes associated with the chronic disease and related clinical or social factors. Finally, in model 3 there is only a temporary concurrence of the chronic disease and COVID-19 infection, but there is not relationship among them, neither have common causes. To determine which configurations are more frequent and which ones apply to each chronic disease, cohort studies that consider the nature of these relationships are required.

**Figure 4.**
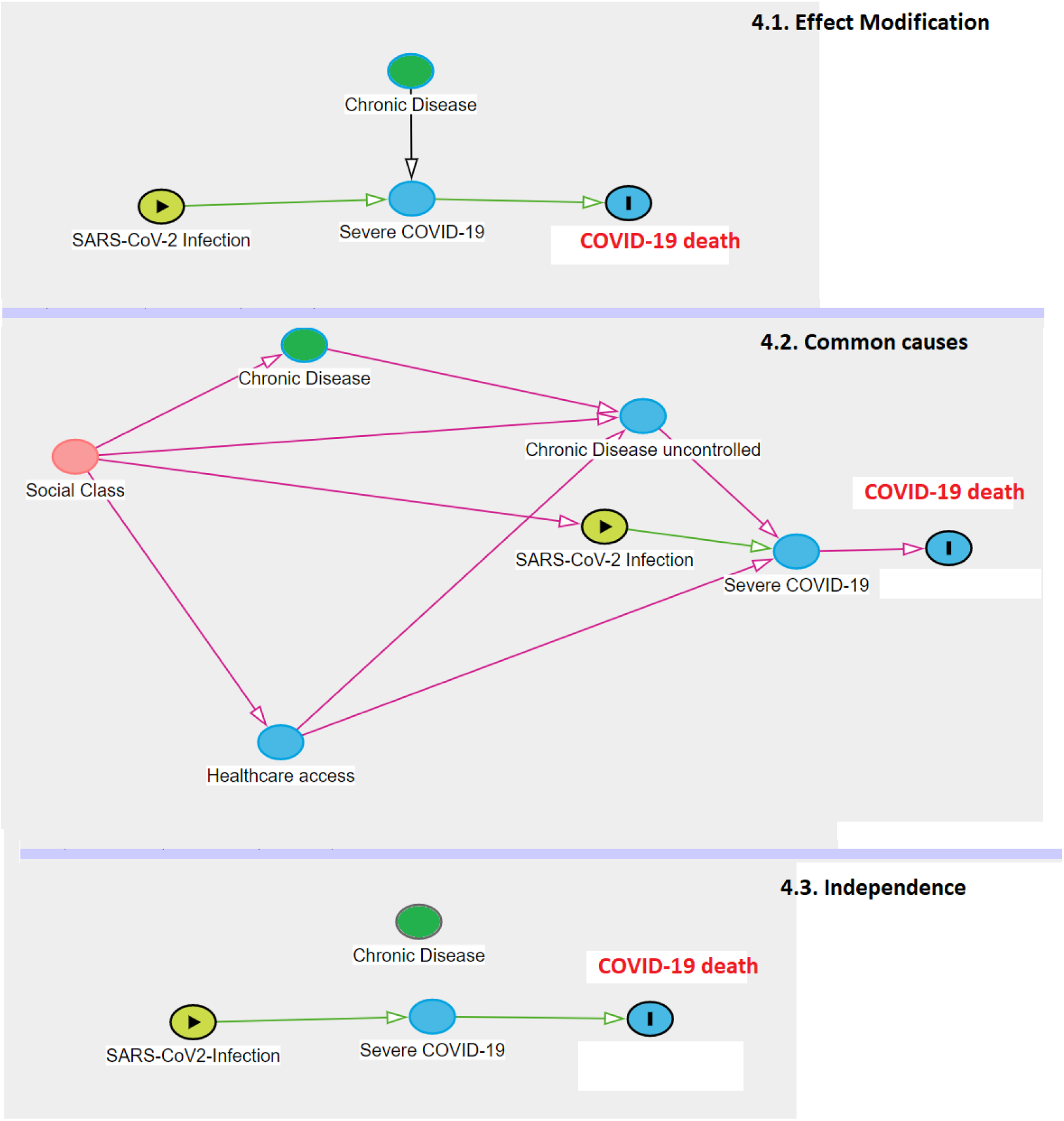
Proposed models between chronic disease and COVID-19 infection observed among Colombian patients.* * Simple theoretical models; in the clinical practice there are many variables involved.

In response to this situation, various specialists have proposed guidelines for managing chronic diseases when it co-occurs with COVID-19. For instance, guidelines for COVID-19 individuals with high blood pressure,^15^ diabetes,^16,17^ cardiovascular disease,^18^ kidney disease,^19^ metabolic and bariatric surgery,^20^ endocrine surgery,^21^ lung cancer,^22^ stroke,^23^ and systemic sclerosis.^24^ There are also recommendations for older adults with multimorbidity and polypharmacy.^25,26^ These types of practices and discussions associated with the proposed changes to manage patients are evidence of the complexity of COVID and the associated multimorbidity. However, the more difficult cases will be individuals with COVID-19 and complex low prevalence diseases.

Multimorbidity should be understood as a vulnerability condition itself, and more than the additive sum of chronic diseases. Multimorbidity is like a concentration of expressions in a unique health-disease process, which means that it’s the joint result of the risk factors and pathophysiological changes associated with each of the health conditions involved. Among older adults, this medical complexity configures the frailty syndrome,^27^ that affects physiological reserves and multiple systems, making older adults more susceptible to complications and deaths from COVID-19, like from other diseases. This could explain the well-recognized relationship between age and severity in COVID-19, which is undoubtedly measured by multimorbidity.

That was the reason why multimorbidity in this study was included as an underlying latent variable in our analysis. Similar experiences with the Patient-Centered Clinical Method (PCCM) questionnaire in Canada, where the complexity of multimorbidity and associated factors can be comparable.^28^ There are even other analyses comparing ways of approaching multimorbidity, which suggest that hierarchical cluster analysis shows comorbidity, while exploratory factor analysis multimorbidity.^29^ Subsequent analyses can use this approach to build scales of risk of death and complications. In addition, diseases that co-occur in time and place configures the existence of a syndemic. For Singer and Clair, when a syndemic occurs is because there are social characteristics that are acting as determinants, so adequate management requires a more comprehensive approach.^27^ In this case, COVID-19 co-occur with different multimorbility patterns, which is specific to a complex emergent disease.

Results presented here should be interpreted taking into account some methodological limitations. First, in this study other conditions that have been used in multimorbidity indices were not available: (osteo)arthritis, depression, hearth failure, depression, osteoporosis, PAVK, vision problems, dementia, hearing problems and angina pectoris^13^. Colombian official database only includes the 10 conditions reported in this study. However, with the exception of heart failure and angina, all other conditions have higher prevalences among older adults. For this reason they would not be directly associated with the risk of becoming infected or dying from COVID-19. Heart failure and angina would be covered by the label “cardiovascular disease”, so we consider that the most relevant morbidities associated with COVID-19 severity were considered.

Another limitation is related to the non-inclusion of socioeconomic health conditions, disability and functional dependence, geriatric syndromes, and mental health problems (mainly depression and dementia). which although their relationship with the probability of death is more direct, if they are theoretically part of multimorbidity from a broader perspective^8^, and could indirectly affect the clinical outcome of the disease, by affecting access to health services, adherence to treatment or its effectiveness, as well as the biological interaction of the underlying conditions. Although the analyses presented here are limited to the patterns of multimorbidity with the highest occurrence, it is important to note that this situation can occur with diseases of lesser occurrence (including neglected diseases) which could be important in some regions. In this case, the medical care becomes a big challenge for clinicians.

In conclusion, in this study the patterns of multimorbidity among people diagnosed with COVID-19 in Colombia during the acute period of infection were presented. Undoubtedly, in the future, more multimorbidity patterns will be known. Similar analyses in different regions that incorporate the different epidemiological patterns^31^ may facilitate the care of people with COVID-19.

## Data Availability

The database can be requested from the Colombian Ministry of Health and Health Protection.

